# Presence of Mediastinal Lymphadenopathy in Hospitalized Covid-19 Patients in a Tertiary Care Hospital in Pakistan – A cross-sectional study

**DOI:** 10.1101/2022.03.10.22272193

**Authors:** Faryal S. Bhatti, Amyn A. Malik, Adeel A. Malik

## Abstract

**Background:** The aim of this study was to investigate the presence of mediastinal lymphadenopathy in hospitalized Covid-19 patients in a tertiary care hospital in the metropolitan city of Lahore, Pakistan from September 2020 till July 2021.

**Methods:** We retrospectively collected data of Covid-19 patients hospitalized from September 2020 till July 2021. Only those patients who tested PCR positive through a nasopharyngeal swab, were enrolled in the study. Patients’ whose data were missing were excluded from this study. Our exclusion criteria included patients who tested negative on Covid-19 PCR, patients with comorbidities that may cause enlarged mediastinal lymphadenopathies such as haemophagocytic lymphohistiocytosis, neoplasia, tuberculosis, sarcoidosis or a systemic disease. The extent of lung involvement in Covid-19 patients was quantified by using a 25-point visual quantitative assessment called the Chest Computed Tomography Score. This score was then correlated with the presence of mediastinal lymphadenopathy.

**Findings:** Of the 210 hospitalized patients included in the study, 131 (62.4%) had mediastinal lymphadenopathy. The mean and median Severity Score of Covid-19 patients with mediastinal lymphadenopathy (mean: 17.1, SD:5.7; median: 17, IQR: 13-23) were higher as compared to those without mediastinal lymphadenopathy (mean: 12.3, SD:5.4; median: 12, IQR:9-16)

**Interpretation:** Our study documents a high prevalence of mediastinal lymphadenopathy in hospitalized patients with Covid-19 with the severity score being higher in its presence representing a more severe course of disease.

## Background

Coronavirus Disease 2019 is the first pandemic to occur since WHO declared H1N1 Influenza a pandemic in 2009.(1-3) Covid-19 is caused by a novel Coronavirus called SARS-CoV-2 (Severe Acute Respiratory Syndrome Coronavirus 2). The main mode of transmission of Covid-19 is through respiratory droplets and via aerosols although transmission can occur through body fluids and from mother to child. Indirect transmission through fomites or contact with objects used on an infected patient can also occur.(4, 5) The clinical features of a Covid-19 infection can range from asymptomatic to mild symptoms of lethargy, fatigue, dry cough and fever to severe symptoms of an acute respiratory distress syndrome.(6)

The standard criterion for diagnosing Covid-19 is by a real-time reverse transcriptase polymerase chain reaction through an oropharyngeal or a nasopharyngeal swab. Sensitivity for rRT-PCR ranges from 60-78%.(7, 8) In cases where lung involvement is suspected a non-enhanced high resolution chest CT scan is performed to supplement a Covid-19 diagnosis and to quantify lung involvement.(7, 9) CT imaging can have a sensitivity of up to 90% but it does have a low specificity. It is difficult to distinguish the viral cause of characteristic abnormalities on CT imaging. However, in PCR confirmed cases of Covid-19, CT imaging is an immensely valuable tool to assess the extent of lung involvement.(9, 10) According to recent studies, mediastinal lymphadenopathy is a commonly reported finding in Covid-19 patients (11-13) although some studies have shown conflicting data as well.(14) Mediastinal lymphadenopathy may be a sign of disease severity. Lymphadenopathy may be attributed to Cytokine Release Syndrome which commonly occurs in critical patients.(14) Currently, the prevalence of mediastinal lymphadenopathy in Covid-19 patients in Pakistan is not known.

To the best of our knowledge, this is the first study in Pakistan to assess the extent of lung involvement in Covid-19 patients using the Chest Computed Tomography Score presented by Li et al.(12) This is a 25-point visual quantitative assessment which has been further categorized into mild [scores 1-16] and severe [scores 17-25].(10) Furthermore, our study correlates the Chest Computed Tomography Score with the presence of mediastinal lymphadenopathy.

## Methods

We conducted this retrospective cross-sectional study at a private, tertiary care hospital in the metropolitan city of Lahore, Pakistan. Lahore is Pakistan’s second largest city with a population of 11.3 million and is the capital of the province of Punjab. It is home to some of Pakistan’s most educated and affluent and is accessible by an international airport with flights from around the world. During the various peaks of the pandemic, Lahore had some of the highest cases of Covid-19 in the country. Our data collection was primarily done at Doctors Hospital & Medical Center, Lahore which is a private hospital where hospital fees are either borne out of pocket or paid for by insurance companies.

### Computed Tomography (CT) Scans

All non-enhanced high-resolution chest CT Scans were performed using a 128 slice CT Scanner (Toshiba Aquilion) with the following parameters: 120 kV, tube voltage 100-200 mAs, rotation time 0·6 s, pitch 1·35. 1 mm slice thickness, sharp convolution kernel reconstructions with a window width of 1500 HU and a window length -500 HU was performed. All images were obtained using a standard dose scanning protocol. The scanning range was from the apex of the lung to the costophrenic angle. Intravenous contrast was not used for these scans. To the best ability of our technical team and the patient’s cooperation to hold his or her breath optimally, the scan was captured in the end-inspiratory phase. The scans were interpreted by in-house qualified radiologists who were blinded to the clinical data.

The high resolution non-enhanced CT were carefully reviewed for distinctive features such as ground glass opacification, consolidations, nodules, reticulation, interlobular septal thickening, crazy paving pattern, linear opacities, subpleural curvilinear line, bronchial wall thickening, mediastinal lymphadenopathy, pleural effusion and pericardial effusion.

### Statistical Analysis

Descriptive analysis was carried out and frequency tables were created to determine distribution of demographic and clinical factors in patients with Covid-19. Prevalence of mediastinal lymphadenopathy was calculated. Chi-square test was used to assess the bivariate association between categorical variables and mediastinal lymphadenopathy. T-test and/or Wilcoxon rank-sum test were used to assess the bivariate association between continuous variables and mediastinal lymphadenopathy. All analysis was carried out on Stata (Version 16, StataCorp LP, College Station, TX, USA).

### Ethical Approval

This study was approved by the Institutional Review Boards (IRB) of Doctors Hospital & Medical Center, Lahore, Pakistan.

## Results

The final sample size for our study was 210 after excluding 40 patients that did not meet the study criteria. Our study population comprised 72.9% (n=153) males and 27.1% (n=57) females. The overall mean age was 59.9 years (SD: 14.5) with median age being 61 (IQR: 50-71; table 1). The mean creatinine (available for 203 patients) was 1.39 (SD: 1.30) while the median creatinine was 1.05 (IQR: 0.86 - 1.33). The mean eGFR (available for 202 patients) was 69.7 (SD: 27.6) while the median eGFR is 71.0 (IQR: 52.2 - 92.6).

**Table 1:**
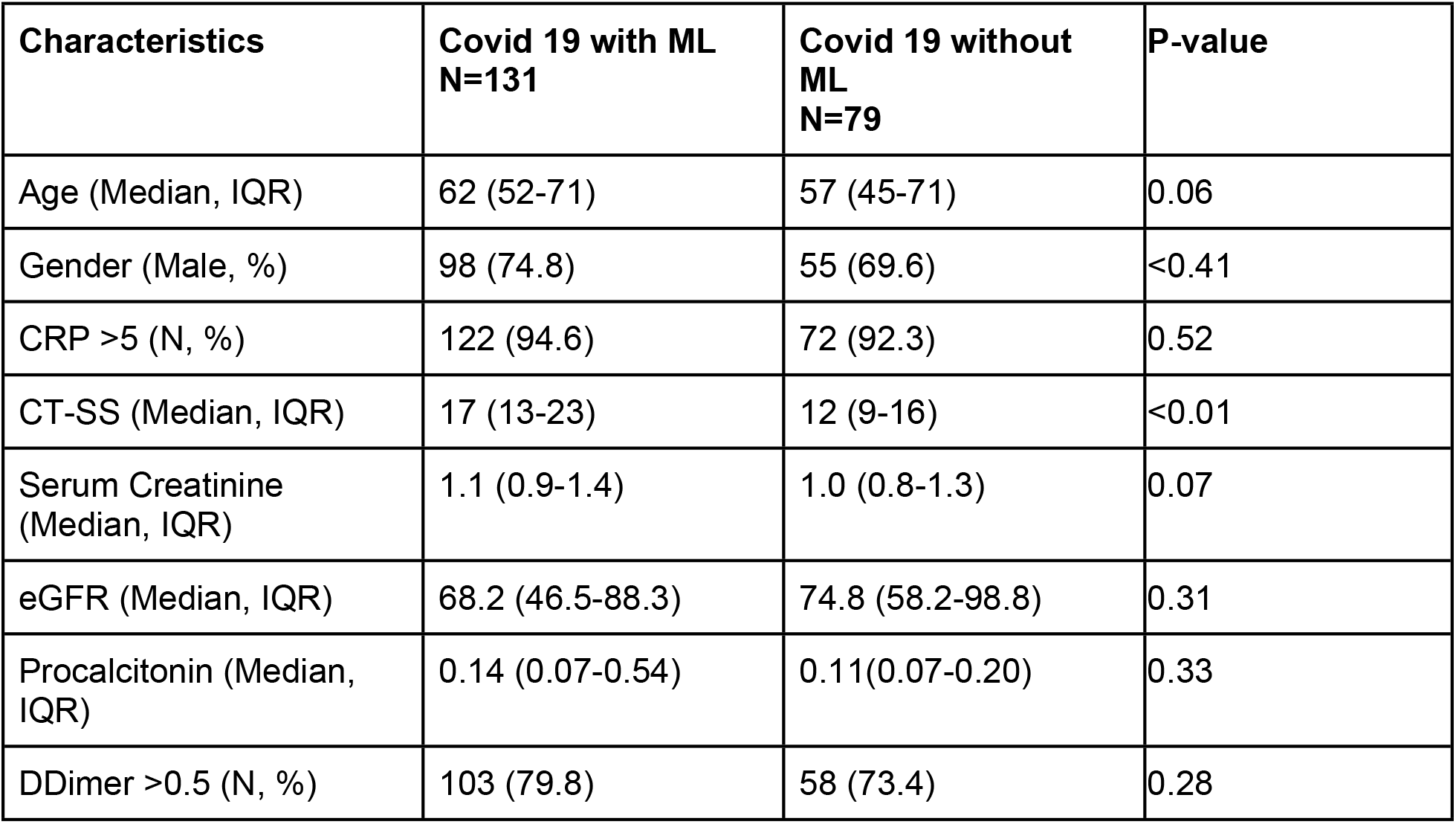
Clinical characteristics of patients with and without mediastinal lymphadenopathy (N=210)

| **Characteristics** | **Covid 19 with ML**<br>**N=131** | **Covid 19 without**<br>**ML**<br>**N=79** | **P-value** |
| --- | --- | --- | --- |
| Age (Median, IQR) | 62 (52-71) | 57 (45-71) | 0.06 |
| Gender (Male, %) | 98 (74.8) | 55 (69.6) | <0.41 |
| CRP >5 (N, %) | 122 (94.6) | 72 (92.3) | 0.52 |
| CT-SS (Median, IQR) | 17 (13-23) | 12 (9-16) | <0.01 |
| Serum Creatinine<br>(Median, IQR) | 1.1 (0.9-1.4) | 1.0 (0.8-1.3) | 0.07 |
| eGFR (Median, IQR) | 68.2 (46.5-88.3) | 74.8 (58.2-98.8) | 0.31 |
| Procalcitonin (Median,<br>IQR) | 0.14 (0.07-0.54) | 0.11(0.07-0.20) | 0.33 |
| DDimer >0.5 (N, %) | 103 (79.8) | 58 (73.4) | 0.28 |

Figures 1-3 show annotated CT with mediastinal lymphadenopathy from a few patients. The prevalence of mediastinal lymphadenopathy was 62.4% (n=131). There was no statistical difference in the gender distribution or ages between those with and without mediastinal lymphadenopathy (table 1).

**Figure 1:**
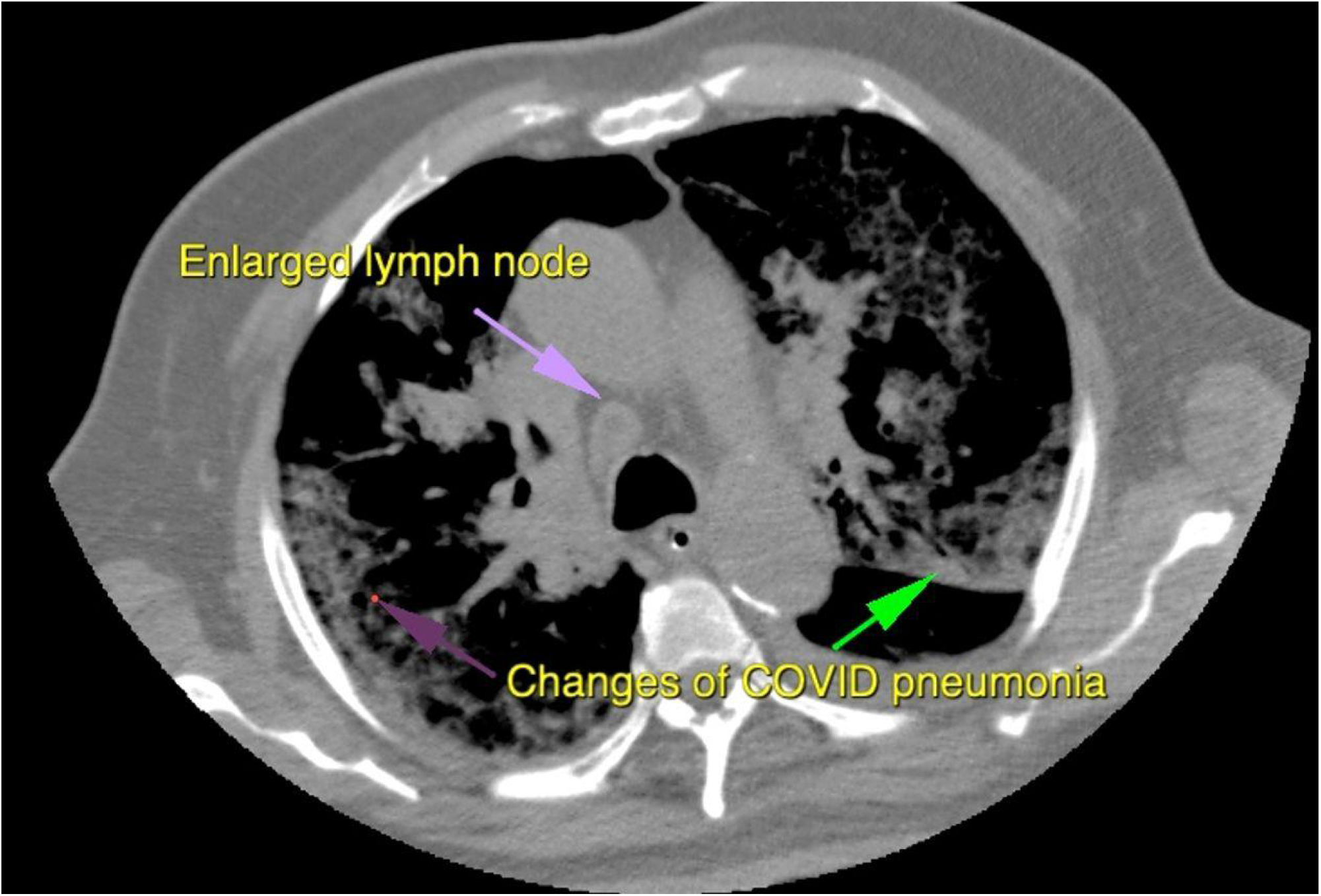
The figure shows an axial CT image with window adjusted to display both enlarged mediastinal lymph nodes along with predominantly sub-pleural ground glass infiltrates typical of Covid-19 Pneumonia.

**Figure 2:**
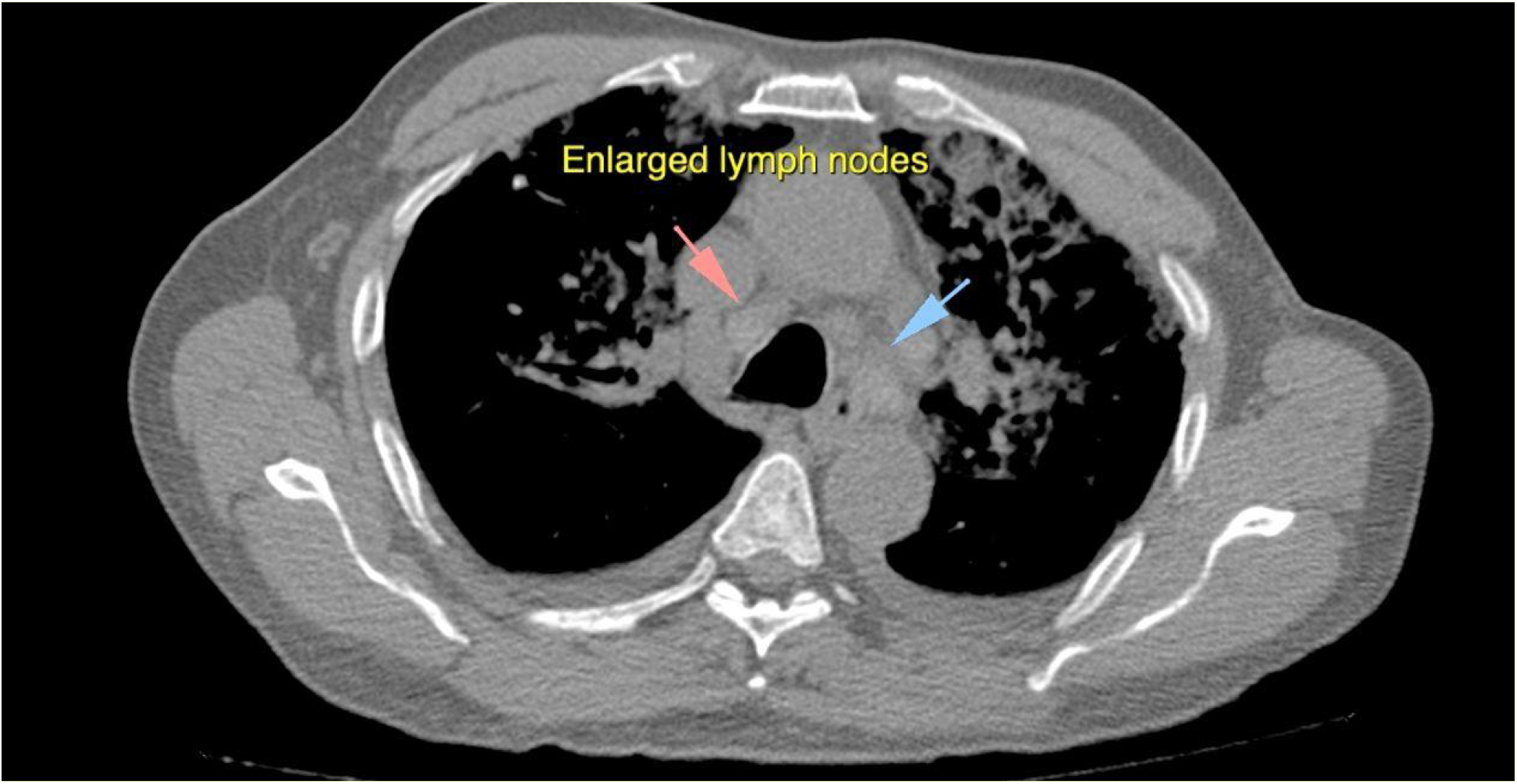
The figure shows Axial CT mediastinal window image showing enlarged mediastinal lymph nodes along with infiltrates in the lungs

**Figure 3:**
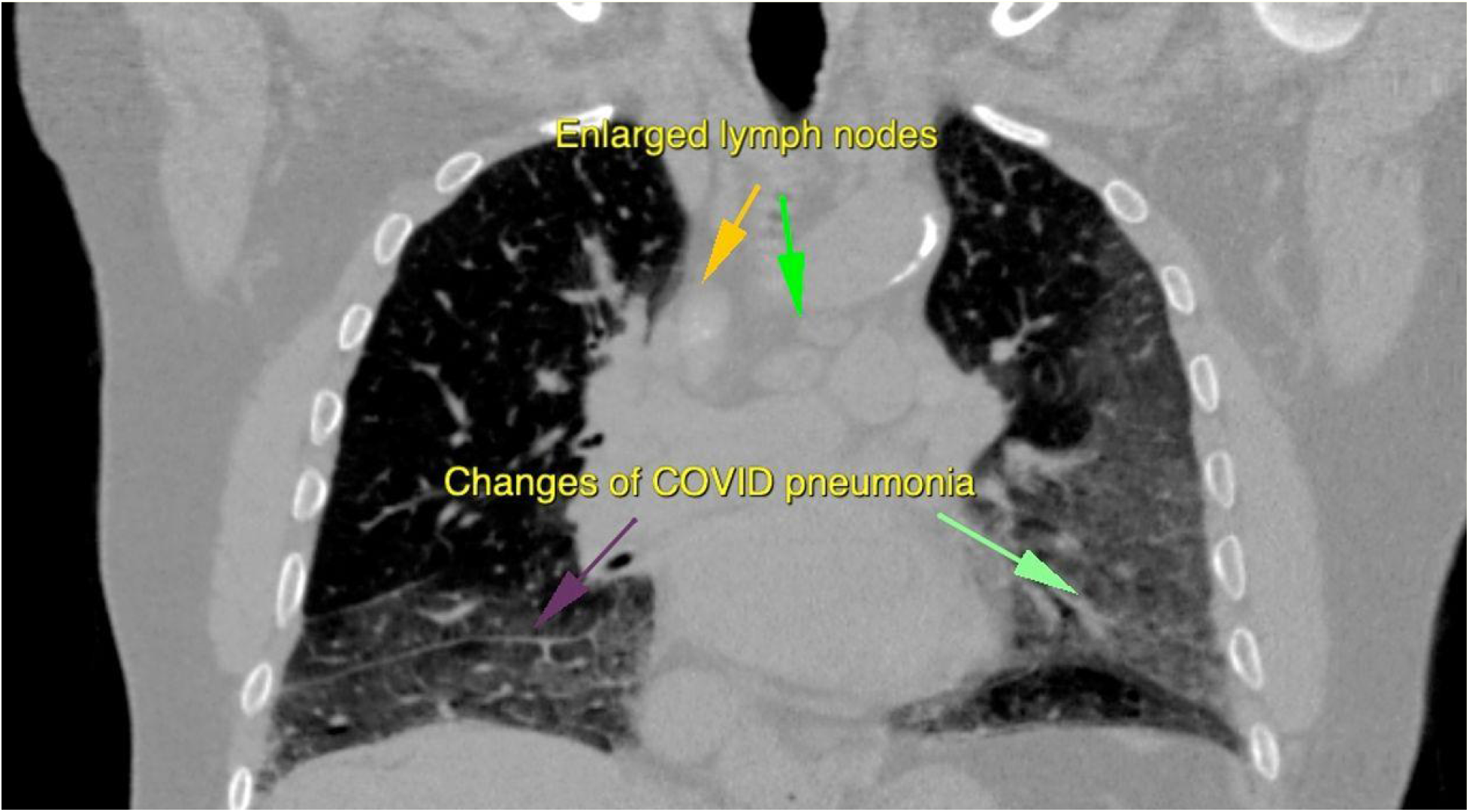
The figure shows a coronal image of CT Scan showing multiple enlarged mediastinal lymph nodes along with confluent patches of ground glass haze suggestive of changes of Covid-19 Pneumonia.

The mean 25-point CT Chest Severity Score for Covid-19 Pneumonia was 15.3 (SD: 6.0) with the median score being 15 (IQR: 11-20). The mean and median Severity Score of Covid-19 patients with mediastinal lymphadenopathy (mean: 17.1, SD:5.7; median: 17, IQR: 13-23) were higher as compared to those without mediastinal lymphadenopathy (mean: 12.3, SD:5.4; median: 12, IQR:9-16; table 1). There was no difference in proportion of patients with elevated procalcitonin, DDimer and CRP between those with and without mediastinal lymphadenopathy. The mean Creatinine between those with mediastinal lymphadenopathy (mean: 1.5, SD: 1.5) and without mediastinal lymphadenopathy (mean: 1.1, SD: 0.7) were statistically different while the median was similar. The mean eGFR between those with mediastinal lymphadenopathy (mean: 65.6, SD: 27.7) and without mediastinal lymphadenopathy (mean: 76.4, SD: 26.4) were statistically different while the median was similar.

## Discussion

Our study aimed to demonstrate the prevalence and significance of mediastinal lymphadenopathy in hospitalized Covid-19 patients in a tertiary care setting in Pakistan. Notably, our study shows that 62.4% of hospitalized Covid-19 patients have mediastinal lymphadenopathy. To be considered as mediastinal lymphadenopathy, the lymph nodes need to be at least 10 mm in short axis. Mediastinal lymphadenopathy can occur as a result of malignant or benign etiologies such as infection or inflammation.(15) As early as February, 2020 a case report in Italy reported the presence of mediastinal lymphadenopathy in 2 patients with Covid-19 who’d traveled from Wuhan, China.(16) A recent literature review of 19 articles (1,155 patients) showed that mediastinal lymphadenopathy was found in 0%-66% of Covid-19 patients. In pediatric patients this number was significantly lower; 0%-8.1%.(17, 18) Similarly, no mediastinal lymphadenopathy was reported in pregnant patients.(19)

A French study involving 15 patients admitted into the ICU reported a 66% prevalence of mediastinal lymphadenopathy. An Italian retrospective study involving hospitalized Covid-19 patients reported a 54.8% prevalence of mediastinal lymphadenopathy.(20) These findings are similar to our study in terms of prevalence of mediastinal lymphadenopathy. A retrospective Chinese study found a 43.5% prevalence of mediastinal lymphadenopathy with Covid-19. However, it is not clear whether the study population included hospitalized patients or not.

In contrast, another Italian retrospective study reviewed Covid-19 patients at Emergency Department admissions and reported a mediastinal lymphadenopathy prevalence of 19% which is significantly lower than what we reported.(21)

Hence, our results are similar to those shown by studies involving hospitalized Covid-19 patients with severe symptoms as indicated by their need for hospitalization especially in an intensive care setting, invasive ventilation, increased oxygen requirement, duration of hospital stay and characteristic features of severity on CT chest and laboratory parameters such as leukopenia and raised inflammatory markers.

Our study reported a significantly higher percentage of males 72.9% hospitalized with Covid-19 as compared to females 27.1%. An Italian retrospective study reported a similar percentage of males 70.2% in their study population of Emergency Department admissions for Covid-19.(21) We did not find a statistical gender difference between patients with mediastinal lymphadenopathy and those without similar to reported by Sardanelli et al.(21) Important to note is that our study reported a higher 25-point CT-Chest Severity Score in Covid-19 patients with mediastinal lymphadenopathy as compared to those without. A retrospective cross-sectional study conducted in Turkey showed that Covid-19 patients with mediastinal lymphadenopathy were significantly older and more likely to have at least one comorbidity along with a higher level of CRP. However, there was no difference in ferritin and procalcitonin levels in both groups. In our study there was no difference in the proportion of patients with elevated procalcitonin, D-Dimer and CRP in Covid-19 patients with and without mediastinal lymphadenopathy. Importantly the Turkish study showed that the presence of mediastinal lymphadenopathy was independently reported to have increased 30-day mortality.(22) Similarly, Sardenelli et al. found that mediastinal lymphadenopathy was more frequently found in Covid-19 patients who died during hospitalization than in those who were discharged.(21)

Kunhua et al. showed that critical Covid-19 patients with high CT scores demonstrated a higher incidence of mediastinal lymphadenopathy.(23) Similarly, a retrospective study of 189 patients in Germany reported lymph node enlargement in around half of their patients with critical disease. The volumetric expansion of lymph nodes may be attributed to a cytokine release syndrome (CRS), a common occurrence in critical patients.(24)

Due to the retrospective nature of our study, we were unable to perform longitudinal assessment of non-enhanced high resolution CT scans. We did not follow how the mediastinal lymphadenopathy progressed over time in different patients. Since we didn’t have previous CT scans for comparison, we had no way of knowing whether the patients comprising our study population didn’t already have mediastinal lymphadenopathy before becoming ill with Covid-19. Smoking is a known contributor of mediastinal lymphadenopathy and data on smoking was not available. Another important consideration is that we did not perform invasive microbiological sampling so coexisting bacterial, fungal and mycobacterial infections could not be ruled out. No biopsies were performed to rule out other causes of mediastinal lymphadenopathy such as malignancies and Sarcoidosis. Lastly, Covid-19 patients with significant findings on non-enhanced high resolution CT chest scans but tested negative on RT-PCR were excluded from our study.

More studies need to be carried out to study the presence of mediastinal lymphadenopathy in hospitalized Covid-19 patients as a prognostic factor. In the meantime, clinicians should be more wary about the presence of mediastinal lymphadenopathy and may want to focus on the timely use of steroids, step up to an intensive care setting, non-invasive ventilation, mechanical ventilation, antibiotics for superadded bacterial infection and early use of monoclonal antibodies.

To the best of our knowledge this is the first study conducted on hospitalized Covid-19 patients in Pakistan to evaluate the presence of mediastinal lymphadenopathy. All our patients tested positive on RT-PCR and had a high resolution non-enhancing CT chest done. Our stringent exclusion criteria ensured that we excluded other causes of mediastinal lymphadenopathy such as fungal infections and tuberculosis.

## Data Availability

Patient records cannot be publicly shared due to patient confidentiality. De-identifie data and specified variables will be made available on request from the corresponding author and the ethics committee of Doctors Hospital & Medical Centre

## Authors’ contribution

FSB conceptualized the study and wrote the protocol; FSB and Amyn AM collected data under supervision from Adeel AM; FSB and Amyn AM performed and reviewed the analysis; FSB and Amyn AM wrote the initial draft of the manuscript. All authors helped interpret the findings, read and approved the final version of the manuscript.

## Acknowledgements

We would like to acknowledge the valuable contribution of the Department of Radiology at Doctors Hospital & Medical Center, Lahore without whom this study would not have been possible. A special thank you to Dr. Sameera Amir for her support and guidance.

